# Obstructive Sleep Apnea destabilizes myocardial repolarization homogeneity

**DOI:** 10.1101/2020.07.22.20159657

**Authors:** Aleksandra Jarecka-Dobroń, Wojciech Braksator, Paweł Chrom

## Abstract

**Background:** Literature shows that patients with obesity and Obstructive Sleep Apnea (OSA), both occurring independently, are more likely to develop cardiovascular diseases and sudden cardiac death (SCD). Assuming that ventricular depolarization is more stable than repolarization then QT interval parameters may be used for heart muscle repolarization assessment for those groups of patients.

**Methods:** There were 121 patients included in the study, both – women and men, aging from 35-65 with visceral obesity. Only healthy patients were included – the ones who were not treated for any chronic disease, taking QT elongating drugs, or were not treated with Continuous Positive Airway Pressure (CPAP) therapy at that time.

**Results:** OSA was diagnosed in 70 patients (including 41 male). A statistically significant difference for QT interval parameters between OSA positive and negative patients was found (p<0.001 for all variables). No difference for QTc max or for QT interval parameters between groups of patients with different OSA severity degrees was found. A correlation between QTVi2 and QTVi3 (p=0.008) and between polygraphy specific parameters and QTc max, QTVi2 and QTVi3 was found.

**Conclusions:** OSA has a negative influence on heart muscle repolarization processes by increasing QTV and QTVi values, without such an impact on QTc max interval among patients with visceral obesity. A temporary intensification of OSA causes additional increase of QTVi value. OSA severity degree expressed by specific parameters taken from polygraphic examination prolongs QTc max and increases QTV and QTVi values.

1. What is the key question? Does Obstructive Sleep Apnea destabilize heart muscle repolarization time heterogeneity expressed by QT interval assessment parameters (QTc, QTV, QTVi)?
2. What is the bottom line? Obstructive Sleep Apnea occurrence presents an essential negative influence on heart muscle repolarization time, as heterogeneity by QTV and QTVi values increase, while QTc max interval remains unaffected among patients with visceral obesity.
3. Why to read on? Knowing that obesity and Obstructive Sleep Apnea (occurring separately) increase the risk of sudden cardiac death, cardiovascular risk evaluation becomes an important point for obese, OSA positive patient’s care.

## Introduction

Visceral obesity is a condition in which abnormally high volume of adipose tissue is deposited in the abdominal region, especially inside the peritoneal cavity and between internal organs. According to the World Health Organization (WHO) and the International Diabetes Federation (IDF) definition, it is diagnosed if waist circumference is more than 94 cm (for men) and more than 80 cm (for women). Obstructive Sleep Apnea (OSA), which occurrence is closely linked to obesity, is characterized by numerous episodes of apnea and hypopnea during sleep caused by temporal, partial or total, upper airways obstruction and it is recognized as an independent cardiovascular risk (1, 2). Obesity has been linked to heart muscle repolarization disorders expressed as QT interval elongation (3). A provocative influence of OSA on heart rhythm disturbances has been frequently researched in the past decade (4, 5). However, only a few reports in the literature mention that OSA may be connected with acquired QT interval elongation (5). Moreover, observations of Sudden Cardiac Death (SCD) increased risk among OSA patients have been described in literature since 1970.

It has been commonly accepted to regard QT interval as an estimation of heart muscle repolarization. QT interval elongation may provoke life threatening ventricular arrhythmias and SCD. QT interval duration shows spontaneous, slight fluctuations in every heartbeat cycle. This phenomenon is specified as QT interval variability (QTV). QTV is primarily a parameter that enables variability of repolarization processes assessment. In standard conditions, at stable heart rhythm frequency, QTV value is low. The best prognostic value, beyond the most frequently used maximum QTc, have QTV, QTV index (QTVi) and QTV assessed in Heart Rate Variability (HRV) low frequency (LF) and high frequency (HF) bands (QTV_LF_, QTV_HF_) (6). A wide clinical importance of the abovementioned parameters has been confirmed in numerous studies and their usage is recommended by European Cardiology Society (ESC) (6-15). The problem of dependence between OSA severity degree and QT interval assessment parameters changes has not been analyzed in literature by using any indicator recommended by European Society of Cardiology (ESC).

## Methods

The current study was a prospective, observational clinical trial performed between September 2016 and August 2019 at a single-center institution specializing in treatment of patients with obstructive sleep apnea. The inclusion criteria were as follows: (1) gender: female or male, (2) age 35-65 years old, (3) visceral obesity, (4) lack of acute or chronic diseases that may have an influence on rhythm or conduction disorders, (5) not undergoing Continuous Positive Airway Pressure therapy (CPAP) or taking drugs that have or may have an influence on QT interval duration (according to Credible Meds list (16)), (6) not consuming grapefruits or grapefruit juice for at least 2 weeks before Holter-ECG examination. Patients, who met all inclusion criteria, underwent subject and physical examination, an over-night Holter-ECG, and polygraphy. After getting all results they were checked against the following exclusion criteria: 1) revealing that information about patient’s chronic illness or drugs therapy was obfuscated, revealing increased fasting serum glucose concentration. It was strongly recommended to every patient to visit their GP for further diagnostics, 2) revealing any important deviation in physical examination i.a. blood pressure taken twice at the visit ≥ 140/ ≥ 90 mmHg, 3) revealing, upon Holter ECG examination, tachycardia or too numerous artifacts making QT interval assessment incredible, 4) too short total sleep time (< 6 hours) registered on polygraphy.

Out of 187 consecutive patients meeting the inclusion criteria, 66 patients met at least one exclusion criterion, leaving 121 patients for the study analyses.

Enrolled patients were asked questions about general frame of mind, daytime and nighttime symptoms that may suggest OSA (according to Epworth Sleepiness Scale (17)). All patients were fully physically examined. Neck and waist circumference were measured according to STEPwise Approach to Surveillance (STEPS) by WHO (18). Upon subject examination (snoring and choking feeling during the sleep) a corrected neck circumference was calculated.

Polygraphy was conducted using MED Recorder device by Infoscan company according to AASM guidelines (19). The device registered blood saturation, heart rate, airflow, chest and abdomen movements, body position, snoring and single lead ECG. OSA was diagnosed according to AASM definition (20): in every patient with Respiratory Disturbance Index (RDI) ≥ 5/hour and with concomitant OSA sings (Epworth Sleepiness Scale ≥ 11 pts) or with RDI ≥ 15/hour. The minimum time of analyzed data without artifacts had to last at least 6 h.

A Holter-ECG examination was conducted using DMS 300-3A device by Oxford company suitable for Cardioscan 10 system. Registration was made simultaneously with polygraphy, during the night. Chosen QT interval assessment parameters were evaluated upon partially automatically analyzed fragments of ECG records. Only nighttime ECG records were analyzed due to maximum comparability (similar patient’s physical activity and minimized, because of limited body movements, artifacts). Moreover, the study aimed to assess a heart’s activity simultaneous to sleep breathing disorders. Only ECG strips with constant heart rate, optimally within 50-70/min limits, were chosen due to the minimum impact of such heart rate on QT interval correction formula. QTc data was calculated upon Bazett’s formula. QTV and QTVi were calculated upon Berger’s formula.

Described above group of patients was divided into groups:

1. Patients with visceral obesity without OSA,
2. Patients with visceral obesity with OSA. Group of patients in which OSA was diagnosed was additionally divided into three subgroups, depending on the breathing disorders severity degree:
  a. OSA of 1^st^ degree (RDI ≥ 5 and <15),
  b. OSA of 2^nd^ degree (RDI ≥ 15 and <30),
  c. OSA of 3^rd^ (RDI ≥ 30).

QT interval assessment parameters were divided into groups:

1. assessment of data of patients only with visceral obesity (no OSA) (QTV1, QTVi1),
2. assessment of data of patients with visceral obesity and OSA, data taken from OSA intervals of minimum severity or from intervals without breathing disorders (QTV2, QTVi2),
3. assessment of data of patients with visceral obesity and OSA, data taken from OSA intervals of maximum severity (QTV3, QTVi3).

Descriptive statistics included means and standard deviations (SDs) (or medians and ranges for non-normally distributed or ordinal data) for continuous variables and frequencies and percentages for categorical variables. Normality assumption was assessed using the Shapiro-Wilk test and homogeneity of variance was assessed using the Levene’s test. For independent variables measured in a continuous scale, the student’s t-test (or the Mann-Whitney test) was used to compare differences between two analyzed groups, while the analysis of variance (ANOVA) (or the Kruskal-Wallis test) was used to compare differences between more than two analyzed groups. In the latter case, the Bonferroni correction (or the Dunn-Bonferroni correction) was used in a *post-hoc* testing. For dependent variables measured in a continuous scale, the paired-sample t-test (or the Wilcoxon test) was used to evaluate differences between measures performed at two timepoints. For independent variables measured in a nominal scale, the chi-squared test (or the Fisher’s exact test) was used to compare differences between two groups. A univariate linear regression was used to assess the influence of RDI on QTc max, QTV and QTVi. Generalized estimating equations (GEE), with identity as a linking function and exchangeable structure of within-group correlation matrix, were used in a multivariate longitudinal analysis covering two timepoints (at no/minimal signs of apnea and at maximum exacerbation of apnea) to evaluate the impact of several clinical factors on QTc max, QTV and QTVi in patients with diagnosed OSA.

## Results

### 1. Clinical characteristic of examined patients

Overall characteristics of the examined population disaggregated into groups with and without OSA, with the analysis of dependence of OSA occurrence on changes in QT interval assessment parameters are presented in Table 1. Population of patients who were diagnosed with OSA numbered 70 people, including 41 men.

**Table 1.**
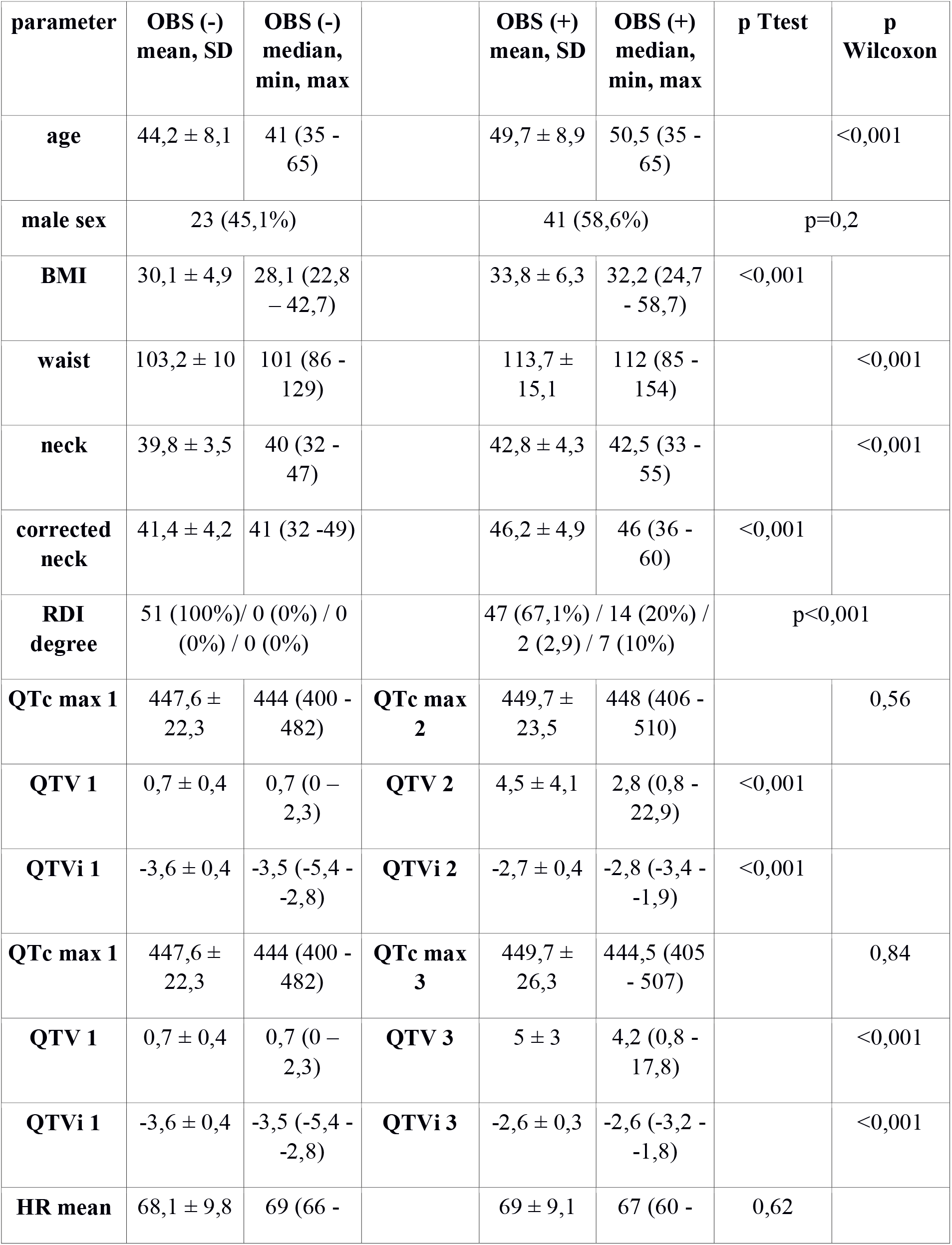
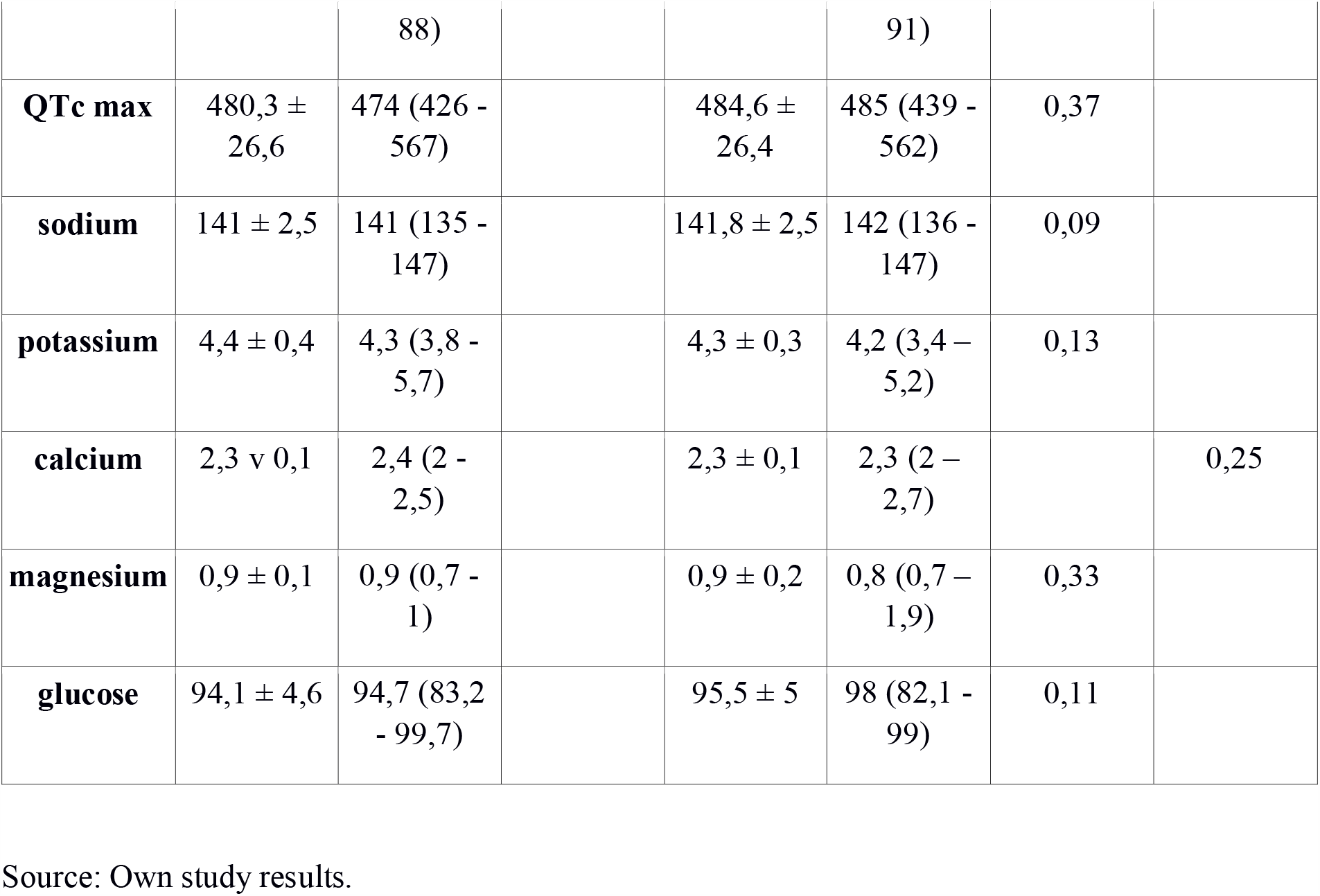
Overall characteristics of population with and without OSA.

### 2. Relationship between OSA occurrence and QT interval

A comparative analysis of QTc max values from the whole Holter-ECG record in OSA positive and OSA negative groups was performed. Arithmetic mean, standard error (SE) and 95% confidence interval (CI) of QTc max are presented in Figure 1. Groups did not differ significantly for QTc max (t= −0.484, degrees of freedom (df) = 119, p=0.630).

**Figure 1.**
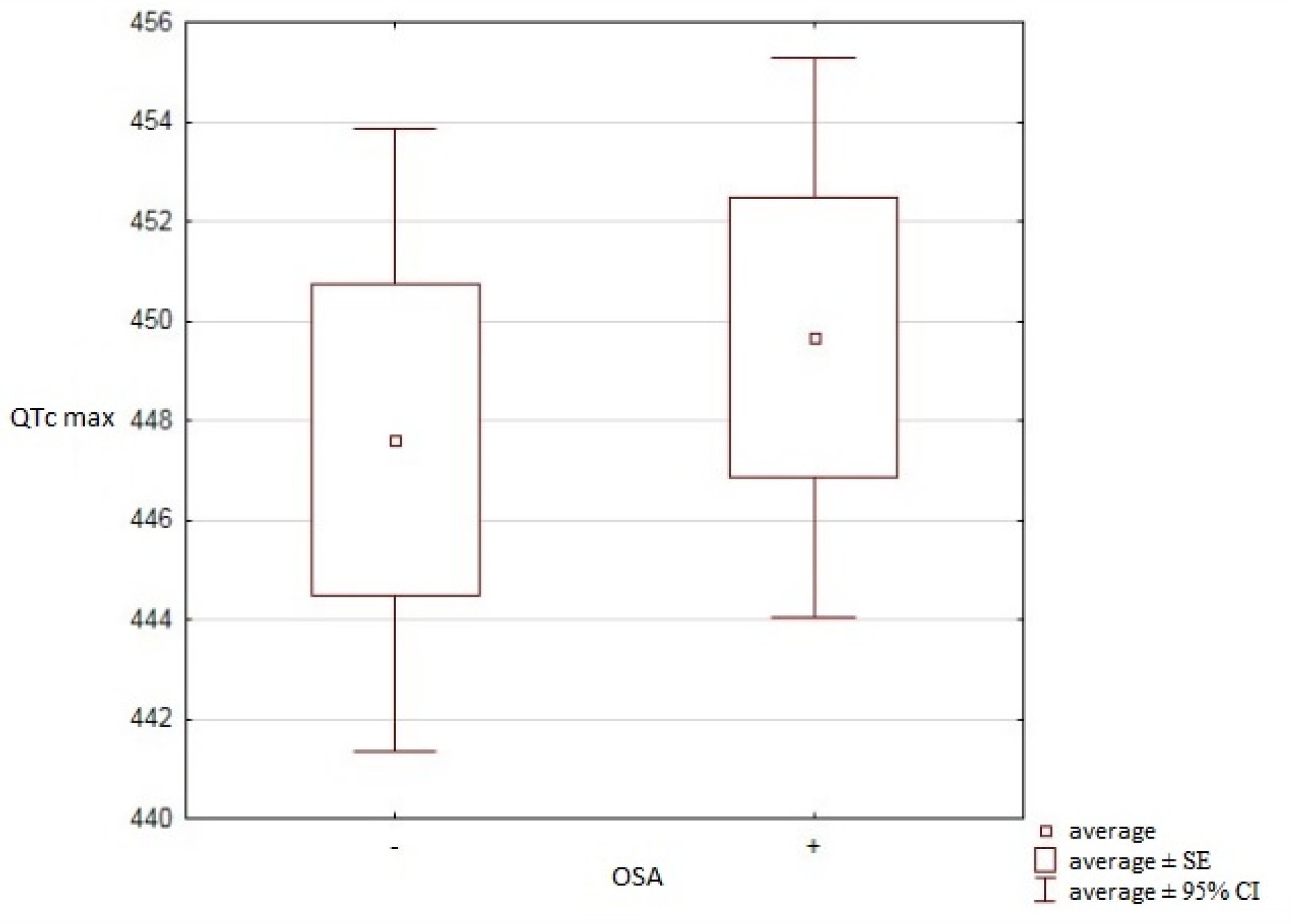
Averages, SE and 95% CI for QTc max in studied OSA positive and negative groups (OSA+ and OSA-). Source: Own study results.

A significant correlation for QTc max 1 on QTc max 2, or for QTc max 1 on QTc max 3 (p=0.6; p=0.84) was not observed. However, a significant correlation for QTV1 on QTV2 as well as for QTV1 on QTV3 was found (p<0.001; p<0.001). A significant correlation for QTVi1 on QTVi2 as well as QTVi1 on QTVi3 was found (p<0.001; p<0.001).

### 3. Relationship between OSA severity degree and QT interval assessment parameters

Analysis results of the correlation between OSA’ severity degree and changes in QT interval assessment parameters are displayed in Table 2.

**Table 2.** A linear regression for a relationship between the QTc max and RDI variables.

| **QTc max 1** | **Coef.** | **Std. Err.** | **t** | **P>\|t\|** | **[95% Conf. Interval]** |  |
| --- | --- | --- | --- | --- | --- | --- |
| RDI | .187138 | .0886293 | 2,11 | 0,037 | .011643 | .3626329 |
| _cons | 445,6936 | 2,526937 | 176,38 | 0,000 | 440,69 | 450,6972 |

Source: Own study results.

A comparative analysis of QTc max values from the whole Holter-ECG record between patient groups with 1^st^, 2^nd^ and 3^rd^ severity degree was performed. Arithmetic mean, SE and 95% CI QTc max are presented in Figure 2. No significant difference in QTc max values of each group was shown (F = 1.355, df = 2, p = 0.265).

**Figure 2.**
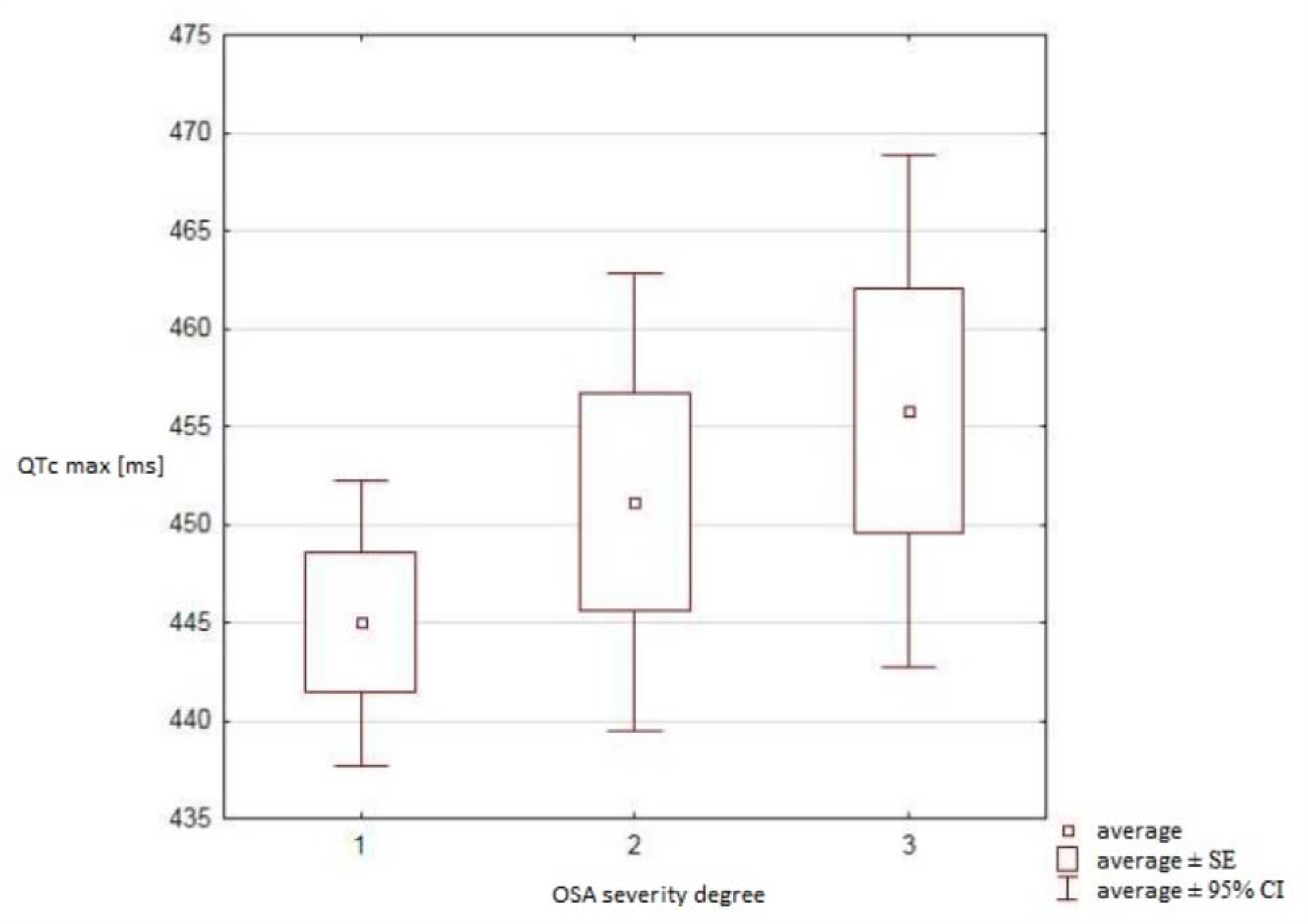
Arithmetic averages, SE and 95% CI for QTC max between patients with OSA of 1^st^, 2^nd^ and 3^rd^ severity degree. Source: Own study results.

No significant difference was found for QTc max taken from ECG record with breathing disorders of a minimum severity or without them (QTc max2) between groups of different OSA severity degrees (p=0.260). No significant difference was found for QTc max taken from ECG record with breathing disorders of a maximum severity (QTc max3) between groups of different OSA severity degrees (p=0.430).

A univariate analysis for RDI influence on QTC max taken from the whole Holter-ECG record assessment was performed. A scatter plot with RDI variable on the X axis and QTc max variable on the Y axis are presented on Figure 3. Regression results with QTc max variable as a dependent variable and RDI variable as an independent variable are shown in the Table 2. It was demonstrated within the regression that RDI increase by 1 per hour causes a statistically significant QTc max elongation by 0.187 ms.

**Figure 3.**
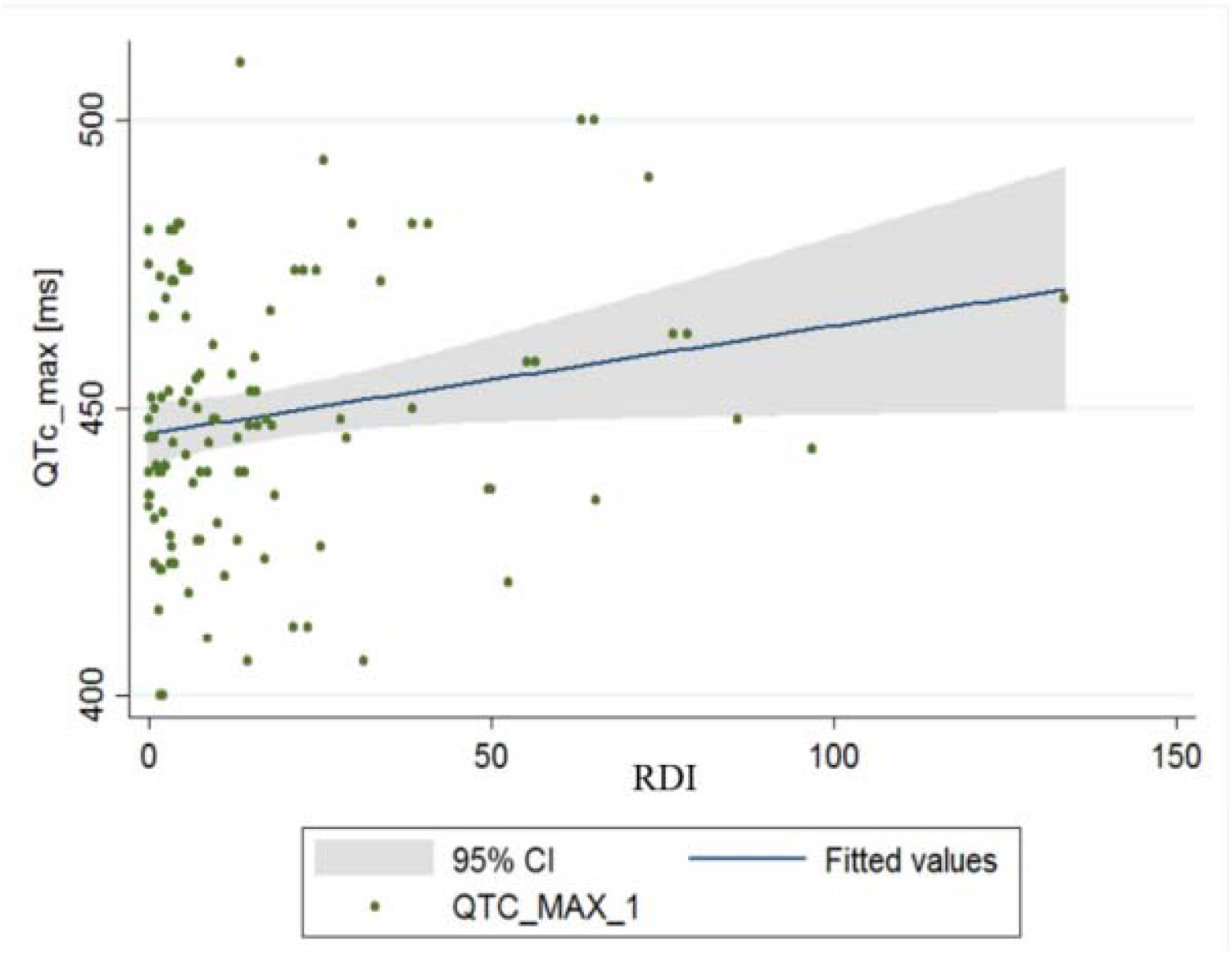
A scatter chart presenting a relationship between the QTc max and RDI variables. Source: Own study results.

No significant difference was found for QTV taken from ECG records with breathing disorders of a minimum severity or without them (QTV2) between groups of different OSA severity degrees (p=0.270). No significant difference was found for QTV taken from ECG records with breathing disorders of a maximum severity (QTV3) between groups of different OSA severity degrees (p=0.10).

A single factor analysis for RDI influence on QTV assessment was conducted. Regression results with QTV variable as a dependent variable and RDI variable as an independent variable are presented in Table 3. It is visible within the regression that RDI increase by 1 per hour causes a statistically significant QTV increase by 0.039 ms.

**Table 3.** A linear regression results describing a relationship between QTV and RDI variables.

| **QTV 1** | **Coef.** | **Std. Err.** | **t** | **P>\|t\|** | **[95% Conf. Interval]** |  |
| --- | --- | --- | --- | --- | --- | --- |
| RDI | .038734 | .0140009 | 2,77 | 0,007 | .0110109 | .0664571 |
| _cons | 2,2877737 | .3991824 | 5,73 | 0,000 | 1,497316 | 3,078158 |

Source: Own study results.

No significant difference was found for QTVi taken from ECG record with breathing disorders of a minimum severity or without them (QTVi 2) between groups of different OSA severity degrees (p=0.150). No significant difference was found for QTVi taken from ECG records with breathing disorders of a maximum severity (QTVi 3) between groups of different OSA severity degrees (p=0.970).

A univariate analysis for RDI influence on QTVi assessment was conducted. Regression results with QTVi variable as a dependent variable and RDI variable as an independent variable are presented in Table 4. It was demonstrated within the regression that RDI increase by 1 per hour causes a statistically significant QTVi increase by 0.011 ms.

**Table 4.**
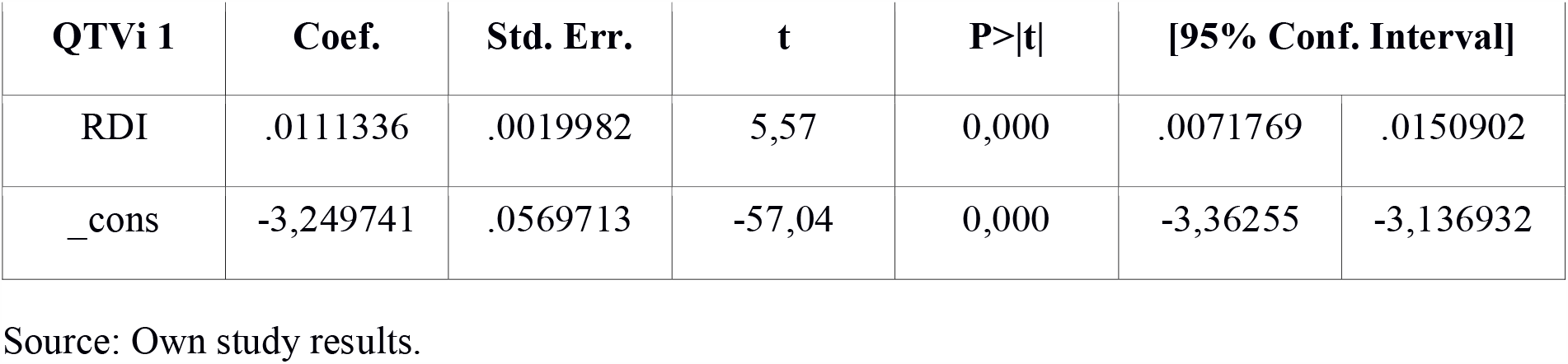
A linear regression results describing a relationship between QTVi and RDI variables.

| QTVi 1 | Coef. | Std. Err. | t | P>\|t\| | [95% Conf. Interval] |  |
| --- | --- | --- | --- | --- | --- | --- |
| RDI | .0111336 | .0019982 | 5,57 | 0,000 | .0071769 | .0150902 |
| _cons | -3,249741 | .0569713 | -57,04 | 0,000 | -3,36255 | -3,136932 |

Source: Own study results.

Comparative characteristics of groups with different OSA severity degrees is presented in Table 5.

**Table 5.** Comparative characteristics of groups with different OSA’s severity degree.

|  | RDI 1,<br>mean,<br>SD | RDI 1,<br>median,<br>min,<br>max | RDI 2,<br>mean,<br>SD | RDI 2,<br>median,<br>min,<br>max | RDI 3,<br>mean,<br>SD | RDI 3,<br>median,<br>min,<br>max | pANOVA | pKruskal-Walis |
| --- | --- | --- | --- | --- | --- | --- | --- | --- |
| **SpO2**<br>**min** | 83 ± 3,5 | 84 (74 -<br>89) | 81,1 ±<br>3,1 | 80,5 (76 -<br>87) | 72,2 ±<br>8,8 | 72 (56 -<br>84) | <0,001 |  |
| **SpO2 av** | 92,8 ±<br>1,7 | 93,2 (89 -<br>95,5) | 92,2 ±<br>1,7 | 92,6<br>(88,8 -<br>95,8) | 90,1 ±<br>4,8 | 91,9<br>(75,2 -<br>94,9) | 0,009 |  |
| **ODI4%** | 9,8 ± 1,7 | 8,6 (4 -<br>22,4) | 26,3 ±<br>16,9 | 22,5<br>(11,7 -<br>89,8) | 62,9 ±<br>28,4 | 52,4<br>(21,2 -<br>132,5) | <0,001 |  |
| **T90** | 7,3 ±<br>12,9 | 1,4 (0 -<br>53,1) | 10,3 ±<br>15,6 | 5,7 (0,4 -<br>65,6) | 28 ± 27,3 | 17,9 (1,4<br>- 89,2) |  | <0,001 |
| **RDI** | 9,3 ± 3,2 | 8,7 (5,2 -<br>14,8) | 21,5 ±<br>4,7 | 21,3<br>(15,6 -<br>29,8) | 60,9 ±<br>25,3 | 55,9<br>(31,4 -<br>133,6) |  | <0,001 |
| **AI** | 0,4 ± 0,7 | 0 (0 -<br>3,2) | 3,7 ± 4,4 | 2 (0 -<br>16,8) | 17,1 ±<br>21,5 | 8 (0 -<br>82,2) |  | <0,001 |
| **QTc max**<br>**2** | 445 ±<br>20,2 | 446 (406<br>- 510) | 451,1 ±<br>23,5 | 448 (412<br>- 493) | 455,8 ±<br>27,9 | 458 (406<br>- 500) | 0,26 |  |
| **QTV 2** | 5,1 ± 5,3 | 2,8 (0,8 -<br>22,9) | 3,2 ± 1,8 | 2,7 (0,8 -<br>7,5) | 4,8 ± 3,5 | 3,1 (1,1 -<br>15) |  | 0,27 |
| **QTVi 2** | -2,8 ± 0,4 | -2,8 (-3,3<br>- -1,9) | -2,8 ± 0,2 | -2,8 (-3,4<br>- -2,4) | -2,8 ± 0,4 | -2,6 (-3,4<br>- -1,9) | 0,15 |  |
| **QTc max 3** | 447,4 ± 26,7 | 436 (405 - 502) | 448,4 ± 25 | 445 (406 - 479) | 454,6 ± 27,6 | 452,45 (405 - 507) |  | 0,43 |
| **QTV 3** | 4,9 ± 3,2 | 4,2 ((0,8 - 17,8) | 4,4 ± 2,8 | 3,4 (1,7 - 12,2) | 59 ± 2,8 | 6,1 (2 - 12,7) |  | 0,1 |
| **QTVi 3** | -2,6 ± 0,3 | -2,6 (-3,2 - -1,8) | -2,6 ± 0,3 | -2,6 (03 - -2) | 02,6 ± 0,3 | -2,6 (-3,2 - -2) | 0,97 |  |
| **HR mean** | 67,1 ± 9,1 | 66,5 (50 - 90) | 69,1 ± 7,9 | 69,5 (53 - 86) | 71,8 ± 9,6 | 68,5 (61 - 91) |  | 0,3 |
| **QTc max** | 482,3 ± 26,3 | 477 (439 - 555) | 481,9 ± 23,4 | 483,5 (441 - 521) | 490,8 v 29,3 | 489 (448 - 562) | 0,47 |  |
| **Na** | 141,4 ± 2,5 | 142 (136 - 146) | 142,3 ± 2,8 | 142 (137 - 147) | 11,8 ± 2,3 | 142 (138 - 146) | 0,5 |  |
| **K** | 4,4 ± 0,3 | 4,3 (3,8 - 5,2) | 4,2 ± 0,3 | 4,2 (3,6 - 4,8) | 4,2 ± 0,3 | 4,2 (3,4 - 4,8) | 0,3 |  |
| **Ca** | 2,3 ± 0,1 | 2,3 (2 - 2,6) | 2,3 ± 0,1 | 2,3 (2,1 - 2,5) | 2,3 ± 0,2 | 2,3 (2 - 2,7) | 0,36 |  |
| **Mg** | 0,9 ± 0,2 | 0,9 (0,7 - 1,9) | 0,9 ± 0,3 | 0,8 (0,8 - 1,9) | 0,8 ± 0,1 | 0,8 (0,7 - 1) | 0,49 |  |
| **glucose** | 95,2 ± 4,9 | 98 (84 - 99) | 95,9 ± 4,6 | 98 (84 - 99) | 95,5 ± 5,7 | 99 (82,1 - 99) | 0,88 |  |

Source: Own study results.

### 4. Relationship between QT interval and temporary OSA intensification

Analysis results of OSA influence on QT interval assessment parameters depending on a temporary OSA intensification are presented in Table 6.

**Table 6.** Results of QT interval assessment parameters analysis according to a temporary OSA’s intensification.

|  | median | minimum | maximum | p Wilcoxon |
| --- | --- | --- | --- | --- |
| QTc max 2 [ms] | 448 | 406 | 510 | p=0,91 |
| QTc max 3 [ms] | 444 | 405 | 507 |  |
| QTV 2 | 2,83 | 0,85 | 22,91 | p=0,06 |
| QTV 3 | 4,24 | 0,85 | 17,82 |  |
| QTVi 2 | -2,77 | -3,38 | -1,87 | p=0,008 |
| QTVi 3 | -2,57 | -3,23 | -1,77 |  |

Source: Own study results.

No significant difference was found for QTc max taken from ECG record with breathing disorders of a minimum severity or without them (QTc max2) and QTc max taken from ECG record with breathing disorders of a maximum severity (QTc max 3) of the same patient (p=0.910).

No significant difference was found for QTV taken from ECG record with breathing disorders of a minimum severity or without them (QTV 2) and QTV taken from ECG record with breathing disorders of a maximum severity (QTV 3) of the same patient (p=0.060).

A significant difference was found for QTVi taken from ECG record with breathing disorders of a minimum severity or without them (QTVi 2) and QTV taken from ECG record with breathing disorders of a maximum severity (QTVi 3) of the same patient (p=0.008).

It is noticeable that a percentage share of male and female sex among groups of patients with and without OSA with a critically elongated QTc max interval diagnosed (for each sex >500 ms) occurs.

The analysis is presented in the Figure 4.

**Figure 4.**
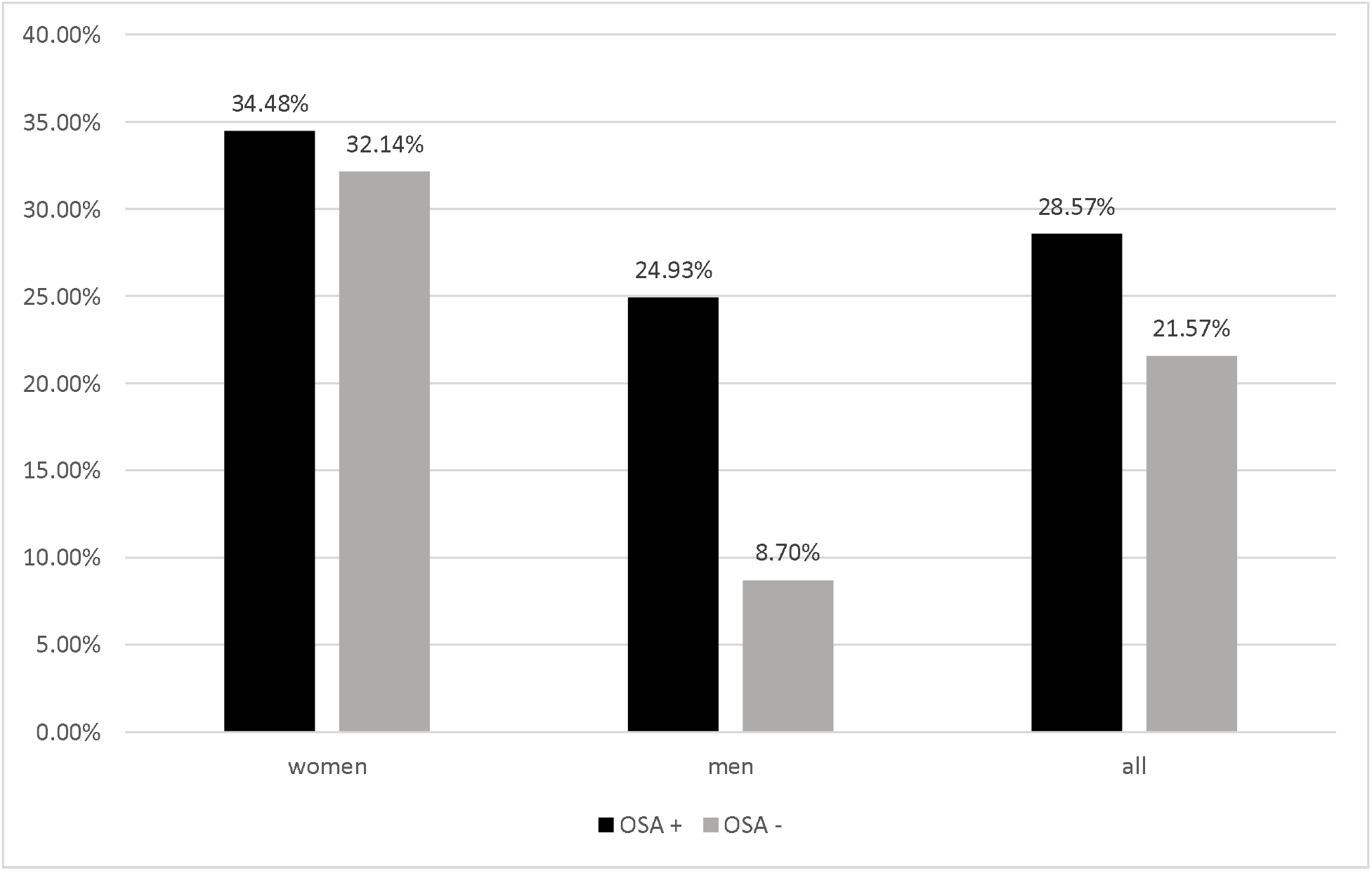
Percentage of patients with critically elongated QTc max interval in studied groups of patients. Source: Own study results.

### 5. Relationship between polygraphic parameters and QT interval

There was a multivariate linear regression conducted for QT interval assessment parameters variability evaluation, taken from the whole overnight Holter-ECG examination and from chosen ECG record fragments, registered simultaneously to breathing disorders of a minimum severity degree or without them as well as from chosen ECG record fragments, registered simultaneously to breathing disorders of a maximum severity degree. Regression assessed variability of those parameters according to variability of chosen polygraphic parameters. With this end in view, three models including variables from polygraphic examination that potentially could interfere with QTc max, QTV and QTVi values, were created. In the first model polygraphic parameters such as: RDI (Respiratory Disturbance Index), AI (Apnea Index), ODI4% (number of episodes of at least 4% saturation decrease), T90 (time of the examination with hypoxia below 90%), SpO_2_ min (minimum registered saturation value), SpO_2_ av (average registered saturation value) with QTc max interval assessment taken from the whole overnight Holter-ECG record were considered. In the second model the same parameters from polygraphic examination with Holter-ECG parameters taken from simultaneous episodes of breathing disturbances of a minimum severity degree or without them (QTc max 2, QTV 2, QTVi 2) were considered. In the third model, above parameters from polygraphic examination with Holter-ECG parameters taken from simultaneous episodes of breathing disturbances of a maximum severity degree (QTc max 3, QTV 3, QTVi 3) were taken into consideration.

A statistically significant difference for the above chosen polygraphic parameters (RDI, ODI4%) and QTc max was found (p=0.004; p=0.005). Also a statistically significant difference for chosen polygraphic parameters (RDI, AI, SpO_2_ min, ODI4%) and QTc max 2 was found (p=0.009; p=0.002; p=0.010). From the same period of time (of minimum or lack of breathing disorders) a statistically significant difference for chosen polygraphic parameters (SpO_2_ min, SpO_2_ av, T90) and QTVi2 was also found (p=0.030; p=0.040; p=0.040). A statistically significant difference for one polygraphic parameter (RDI) and QTc max 3 (p=0.040) as well as for RDI, AI, ODI4% and QTV3 was found (p=0.030; p=0.0300; p=0.02). Moreover, all results appeared to be independent of age and BMI. Analysis results are presented in the Table 7.

**Table 7.**
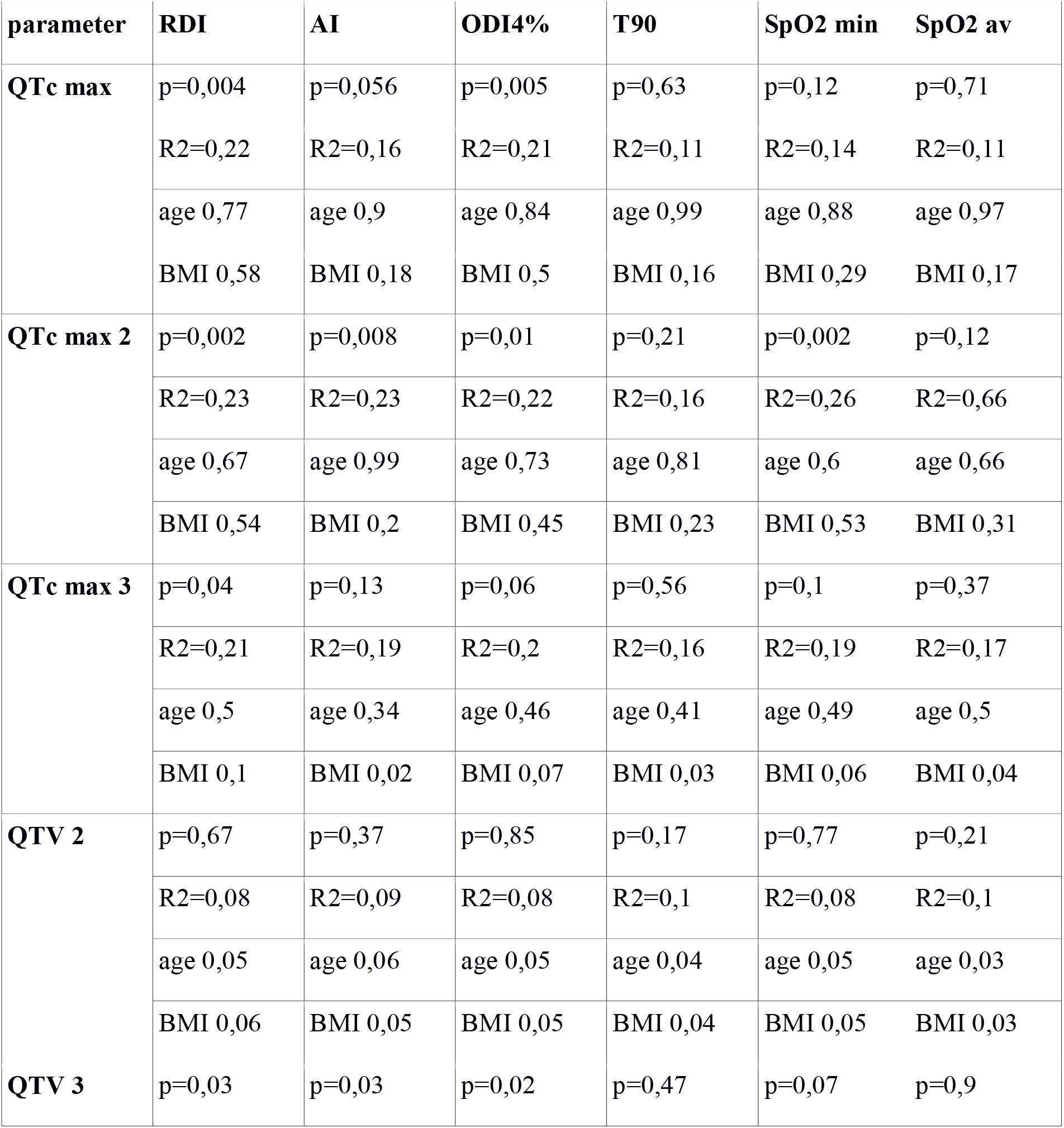
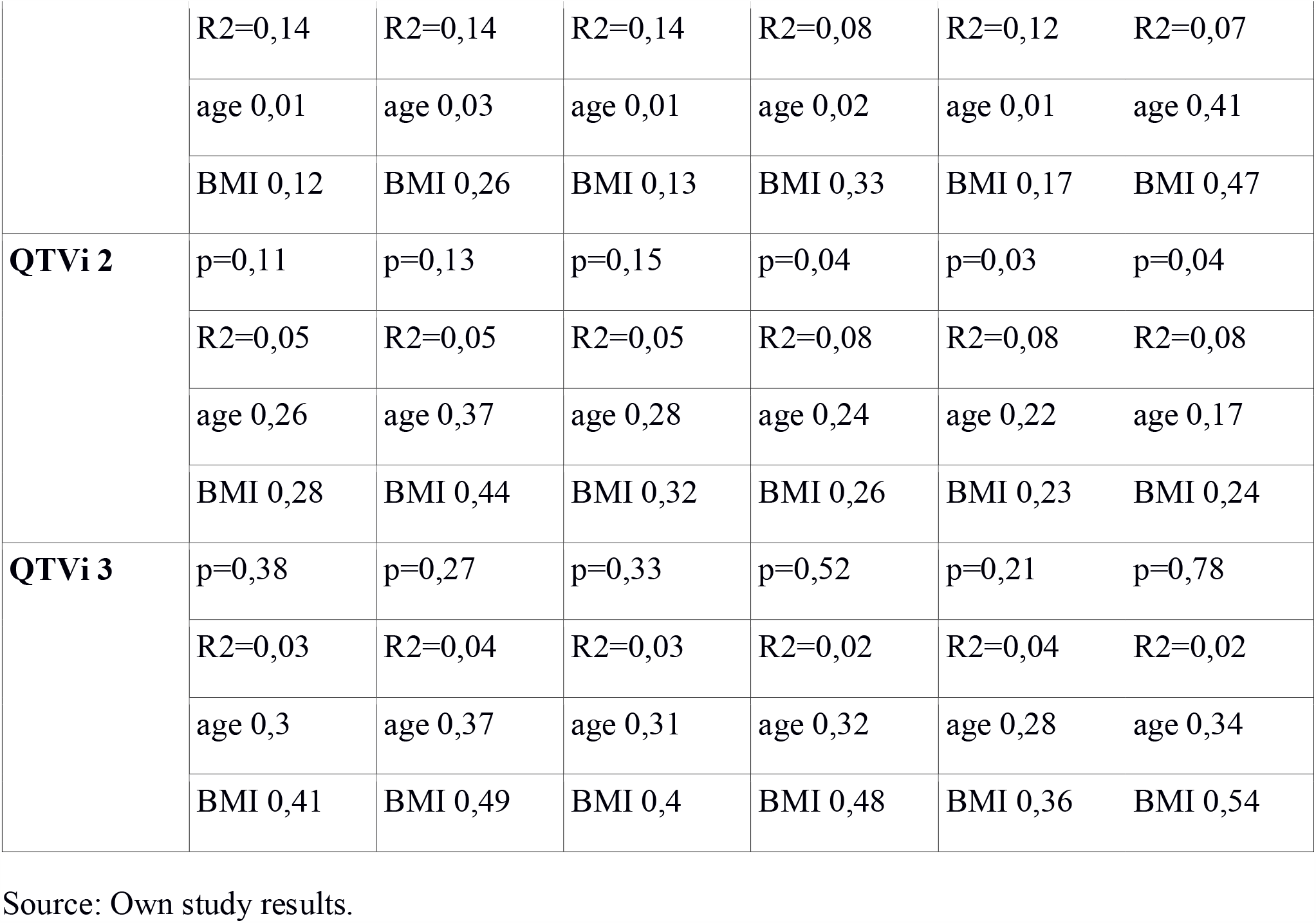
A multifactor regression analysis of polygraphic and QT interval analysis parameters results.

## Discussion

So far only a few studies evaluated the visceral fat tissue’s potential to disturb heart muscles electrophysiological functioning (21-23). Results of those studies, however taken on small groups of patients, may be indicative for a negative influence of biochemical factors released to the bloodstream by visceral fat tissue on the heart muscle. What is more, only a few researchers have decided to study a correlation between OSA and heart muscle repolarization disturbances. An important, pathological relationship between those two diseases was observed by Baumert et al. (24).

All patients had a sinus rhythm of a comparable rate, about 68-69/min. Presented in the Table 1 basic, anthropometric parameters characterizing the studied population are similar to those described by other researchers (21, 22, 25, 26). Significantly more patients with OSA of a severe degree were men. Also, OSA positive patients were statistically older and more obese (BMI, waist and neck circumference). It emphasizes the importance of tests for OSA at least among middle-aged, obese men. Studied groups heterogeneity, in terms of the above mentioned parameters, is a result of the conducted study model. A significant difference in heart rate between OSA positive and negative groups was not shown, which was one of the methodological assumptions – it enabled a reliable assessment of QT and QTc values from Holter-ECG records. Results of so far published studies regarding OSA usually took into account only patients with at least moderate OSA severity degree (most with a severe degree) and did not compare the results with a validation cohort or studied groups were very small.

In the conducted analysis the influence of OSA on QTc interval (taken from the whole overnight Holter-ECG record) elongation was not confirmed. However, the regression model showed that RDI increase by 1 event/hour causes a significant QTc interval elongation. A significant relationship between OSA and increase of QTV and QTVi was confirmed. Also, the regression model confirmed that RDI increase by 1 event/hour causes a statistically significant QTV and QTVi increase. No differences in QT parameters in groups of OSA patients of a different severity degree were shown. Those findings may be indicative of a constant, negative OSA influence on the heart muscle repolarization stability.

Baumert et al in a retrospective study showed a correlation between QTV variance and OSA incidence in at least medium severity degree (24). However, the study group was smaller and even more heterogeneous in terms of age and body weight than in the presented study. Also, Viigimae et al, while examining OSA patients, observed a correlation between breathing disorders and QTV by analyzing different sleep stages (27). So far, there was no study concerning this problem conducted on a bigger than 50 patients group, with no less control group. In the study supervised by Gami data of over 10 000 patients was analyzed; on its base OSA was identified as an independent risk factor for SCD (28). SCD among OSA patients is also more frequently observed at night than during the day, in contrast to the general SCD population (29). The results of the presented study may explain pathophysiology of this phenomenon.

Moreover, a gender percentage participation in groups of patients with and without OSA was assessed, in which a critically elongated QTc max was diagnosed (for both sexes >500 ms). It is shown that a critically elongated QTc max value was diagnosed more often among men with OSA than among men without this disease. A similar regularity was not observed among women. Therefore, it could mean that male gender additionally generates an adverse influence of OSA on heart muscle repolarization. All patients in whom a critically elongated QTc max was diagnosed were referred for further diagnostics.

An interesting result was also produced by the analysis of the influence of OSA temporal intensification on analyzed QT interval assessment parameters. There were 4-5 minutes lasting fragments of ECG records chosen, for which there were simultaneously recorded breathing disorders of minimum severity (or lack of them) for each patient. Subsequently, they were compared with, also 4-5 minutes lasting, fragments of ECG record of episodes of OSA of maximum intensification, for the same patient. A statistically significant difference for QTVi between episodes of minimum and maximum OSA intensification was noticed. It proves that there is an essential adverse influence, of even temporal OSA intensification, on heart muscle repolarization. Therefore, this result may explain results of the above mentioned studies, in which an increase of SCD incidence among OSA patients was shown especially during night time, when breathing disorders occur and get worse. Close to statistical significance also the result of the relationship between QTV calculated from analyzed ECG fragments has been placed. Therefore, a further evaluation of this dependence assessed on a bigger data would be interesting. The results of the above presented analysis may present an additional, next to continuous, negative influence of breathing disorders on heart muscle repolarization, appearing in moments of increased hypoxia. The analysis had a typically pilotage style. There are no similar calculations in available literature, analyzing a correlation between temporal OSA intensification on QT interval change.

Many, significant associations between the most important parameters from polygraphy (i.e. RDI, AI, ODI4%, T90, minimum and average blood saturation) and QT interval assessment parameters were shown. Those associations applied to relationships between both quantitative and qualitative OSA intensification parameters, independently of age or body weight of patients. Those results prove that the number of hypoxia episodes, as well as their depth and duration, have a crucial influence on heart muscle repolarization processes homogeneity. The analysis for so many polygraphic parameters has not been carried out in literature yet.

## Limitations of the study

A relatively small number of examined population and its heterogeneity in terms of age and obesity degree may be perceived as a limitation of the study. Another limiting factor turned out to be the usage of Holter-ECG analyzing software, preventing QTV analysis in HRV of LF and HF band, which made an analysis of QT interval parameters, in the light of QTV assessment guidelines, incomplete (6). The last limiting factor is lack of a long-term observation of the examined population which could help to make more conclusions concerning the clinical significance of observed differences. However, a follow-up study after one and five years for mortality evaluation is planned.

## Data Availability

Participants' enrollment to the clinical trial had begun in September 2016 and had finished in August 2019.

